# Combining structured and unstructured data in eMRs to create clinically-defined eMR-derived cohorts

**DOI:** 10.1101/2020.07.27.20163279

**Authors:** Charmaine S Tam, Janice Gullick, Aldo Saavedra, Stephen T Vernon, Gemma A Figtree, Clara K Chow, Michelle Cretikos, Richard W Morris, Maged William, Jonathan Morris, David Brieger

**Author notes:** **Corresponding author:** Dr Charmaine Tam, Office 543, Level 5, School of Computer Science (J12), The University of Sydney, NSW, 2006 Australia. Email.

## Abstract

**Background:** There have been few studies describing how eMR systems can be systematically queried to identify clinically-defined populations and limited studies utilising free-text in this process. The aim of this study is to provide a generalisable methodology for constructing clinically-defined eMR-derived patient cohorts using structured and unstructured data in eMRs.

**Methods:** Patients with possible acute coronary syndrome (ACS) were used as an exemplar. Cardiologists defined clinical criteria for patients presenting with possible ACS. These were mapped to data tables within the eMR system creating seven inclusion criteria comprised of structured data fields (orders and investigations, procedures, scanned electrocardiogram (ECG) images, and diagnostic codes) and unstructured clinical documentation. Data were extracted from two local health districts (LHD) in Sydney, Australia. Outcome measures included examination of the relative contribution of individual inclusion criteria to the identification of eligible encounters, comparisons between inclusion criterion and evaluation of consistency of data extracts across years and LHDs.

**Results:** Among 802,742 encounters in a 5 year dataset (1/1/13 to 30/12/17), the presence of an ECG image (54.8% of encounters) and symptoms and keywords in clinical documentation (41.4-64.0%) were used most often to identify presentations of possible ACS. Orders and investigations (27.3%) and procedures (1.4%), were less often present for identified presentations. Relevant ICD-10/SNOMED codes were present for 3.7% of identified encounters. Similar trends were seen when the two LHDs were examined separately, and across years.

**Conclusions:** Clinically-defined eMR-derived cohorts combining structured and unstructured data during cohort identification is prerequisite for critical validation work required for secondary use of eMR data.

## INTRODUCTION

The widespread adoption of electronic medical records (eMR) offers unprecedented opportunities to rapidly ascertain and examine clinical data at large-scale and low cost; such information is essential for applications such as audit and feedback, near real-time clinical decision support as well as supporting research objectives through cohort studies, registries and large-scale pragmatic clinical trials [1-3]. Administrative coding systems such as ICD-10 provide a translation of healthcare diagnoses, procedures, medical services, and medical equipment into universal codes primarily used for medical billing. Whilst these are invaluable for their intended purpose-medical billing-they are often used for monitoring population health status and utilisation of healthcare services [4]. Furthermore such diagnostic codes do not provide a granular view of a patient’s presentation, severity of disease and clinical sequence during an episode of care [4-6]. Improved techniques are required to maximise the depth and accuracy of information extracted from the eMR to transform our ability to monitor performance, identify gaps and inequities, and optimise efficiencies of testing new clinical pathways and innovations.

The development of robust methodologies that enable identification of clinical-defined cohorts from the overall patient population captured *within* eMR systems are a critical first step [7]. The most straightforward approach is to use clearly defined events or procedure codes (eg. type of surgery, cancer diagnosis) associated with the hospitalisation to identify cohorts [8, 9]. However, diagnostic codes alone are insufficient for identifying clinical eMR-derived cohorts due to strict rules adhered to for coding, underreporting and the complexity of diseases being assigned a single code [10]. The cohort identification process becomes even more challenging when diseases and conditions have heterogeneous aetiology and a spectrum of severity [11-13].

There have been few reports describing how eMR systems can be systematically queried to identify and reliably extract information from a clinically-defined cohort of interest and limited studies leveraging the >70% of the eMR that is captured in free-text during this process [14-18]. Furthermore, previous studies have used structured data fields (eg. ICD10-code) to identify the population for data extraction [19, 20] or cohort identification has occurred *after* the data has been extracted from the eMR system [10, 21]. In addition, studies which have compared coding to clinician-determined diagnoses report agreement ranging from 80-85% [22, 23].

The aim of this study was to develop a generalisable methodology for creating high-fidelity clinically-derived eMR cohorts for complex diseases/conditions which cannot exclusively use diagnostic or procedure codes to identify a cohort of interest. Patients with possible acute coronary syndrome (ACS) were selected as an exemplar given it’s heterogeneous aetiology and spectrum of severity during presentation. This approach can be applied to other complex diseases/conditions including but not limited to mental illness, asthma, rheumatoid arthritis, chronic kidney disease [11-13, 17].

## METHODS

### Source population

The source population presented to health care facilities in Northern Sydney Local Health District (LHD) and Central Coast LHD, two of the fifteen LHDs in New South Wales (NSW), Australia. LHDs are responsible for managing public hospitals and health institutions and for providing health services within a geographical area. Within Northern Sydney LHD and Central Coast LHD, there were eight publicly-funded hospitals that admitted patients with possible ACS. This included two tertiary hospitals with 24-hour percutaneous coronary intervention (PCI)-capability and six referral hospitals; all of which used Cerner Millennium information systems. These two LHDs had a combined estimated resident population of 1.26 million people [24]. Ethics and governance approval for the study (called *SPEED-EXTRACT*) was provided by the Northern Sydney Local Health District (NSLHD) Human Research Ethics Committee. In this methodology paper, we followed the guidelines developed in the RECORD statement for reporting studies conducted using observational, routinely-collected health data [25].

### Study inclusion criteria and rationale

A multi-disciplinary team consisting of 5 clinicians (cardiologists, population health physicians, nurses) and 5 electronic data experts (data engineers, business analysts, data analysts, analytics translator). In a series of initial meetings, the cardiologists defined clinical criteria to identify patients presenting with possible ACS. The data experts then mapped the clinical criteria to discrete data tables within the Cerner Millenium eMR system to locate the data elements required for identification of patients using these clinical criteria. This mapping process was discussed and agreed upon at iterative meetings over a 3-month period with the multi-disciplinary team. This approach of using a multi-disciplinary team to define clinical criteria which can then be mapped to discrete data tables in the eMR can be applied to any condition/disease of interest. During this process, we considered the previously described phenotype algorithm model workflow model for portable algorithms [26], against existing technological constraints for extracting data from Cerner eMR systems within our health jurisdiction.

We took a liberal approach to the overall design of the inclusion criteria for several reasons. Firstly, we wished to validate ICD-10 coding against expert clinical diagnosis for ST-elevation myocardial infarction (STEMI), one of the main types of ACS. The aim of this was to quantify the extent of misclassification [27] and to obtain a labelled dataset to which machine learning could be applied to accurately identify patients with STEMI. A broad approach was required to identify STEMIs in the eMR extract that had not been identified through routine ICD-10 coding. We also wished to create a comprehensive clinical eMR data platform that could be interrogated for a range of cardiovascular-related questions in the future, and would be inclusive enough to accommodate changes in clinical definitions and guidelines [28, 29]. To this end, seven inclusion criteria to identify possible ACS were developed (Table 1). If any of the seven inclusion criteria were met for an encounter during the study period, the encounter (termed an “*eligible*” encounter) was deemed eligible for inclusion into the study population. An encounter was defined as an electronically recorded interaction between a patient and healthcare provider, characterised by a unique identifier and admission time. Once an encounter met any of the seven inclusion criteria, all clinical, biochemical and demographic information contained within the eMR were extracted. This also included information from “*reference”* encounters for the individual patient dating back to 2002 when the eMR system was first implemented in the health district.

**Table 1.** Study inclusion criteria

| Inclusion Criteria | Definition |
| --- | --- |
| 1 | The “Reason for Visit” (free text field) for the presentation contained any of the ACS-related symptoms or keywords described in Supplementary Information Part 1. |
| 2 | The patient was placed on a cardiac pathway care plan OR the information collected during emergency department triage (free-text), separate to the Presenting Information field above, contained any of the list of ACS-related symptoms or keywords (Supplementary Information Part 1). |
| 3 | Orders were placed in the eMR for a troponin test OR a 12 lead ECG OR for any of the following investigations: coronary angiogram, exercise stress test, stress echocardiogram, sestamibi scan, CT coronary angiogram, CT pulmonary angiogram |
| 4 | The patient’s eMR contained a “Cardiac Monitoring” form, meaning that the patient had been placed on a cardiac monitoring pathway |
| 5 | The patient had a result recorded in the eMR from a sestamibi scan, CT coronary angiogram, CT aortic angiogram or CT pulmonary angiogram |
| 6 | Any of the International Classification of Diseases 10<sup>th</sup> Revision- Australian Modification (ICD10-AM) codes recorded for an encounter started with “I21”, “I22”, “I23”, “I24” or “I25” OR the episode of care had any of the following diagnoses (SNOMED-CT) recorded in the eMR: “acute myocardial infarction”, “acute non-ST segment elevation”, “acute ST segment elevation”, “acute non-q wave infarction”, “angina” |
| 7 | The encounter contained a scanned 12-lead ECG image. |

For criteria (1) and (2), an initial list of ACS-related symptoms, keywords and abbreviations, for patients presenting with possible ACS was developed in consultation with the clinical reference group. The list was extended by manual review of ∼50 ED triage forms in patients with an ICD10 code for STEMI (I21.0, I21.1, I21.2, I21.3) or NSTEMI (I21.4). Next, we examined the frequency of each of the search terms for relevant data fields within a subset of ED triage forms obtained from a 3-month test extract (n∼30,000 encounters between 1/4/17 to 30/6/17). Given the unstructured nature of free-text, we also identified and included misspellings and abbreviations for each of the terms explicitly due to the text processing limitations of Cerner Command Language (CCL) used to extract the data. If the keywords were present at least 20 times in the subset of 30,000 ED Triage forms, they were then included in the final list of search terms of ACS-related symptoms and keywords.

### eMR Data extraction

Data extraction was performed for a single continuous five-year time period between 1/1/13 to 31/12/17. Encounters were extracted through the execution of a bespoke Cerner Command Language (CCL) script which contained seven functions (representing the seven inclusion criteria) which were designed to identify *eligible* encounters. Data extraction was performed by an external party (MKM Health, Chatswood, NSW) and in line with the HREC approval, the study investigators had no direct access to the eMR information systems or contained within the eMR. A visual representation of the data tables extracted in this study is shown in Figure 1. Approaches for ensuring data quality and the operational framework of the study (data management, security and governance) are described in Supplementary Information, Part 2.

**Figure 1.**
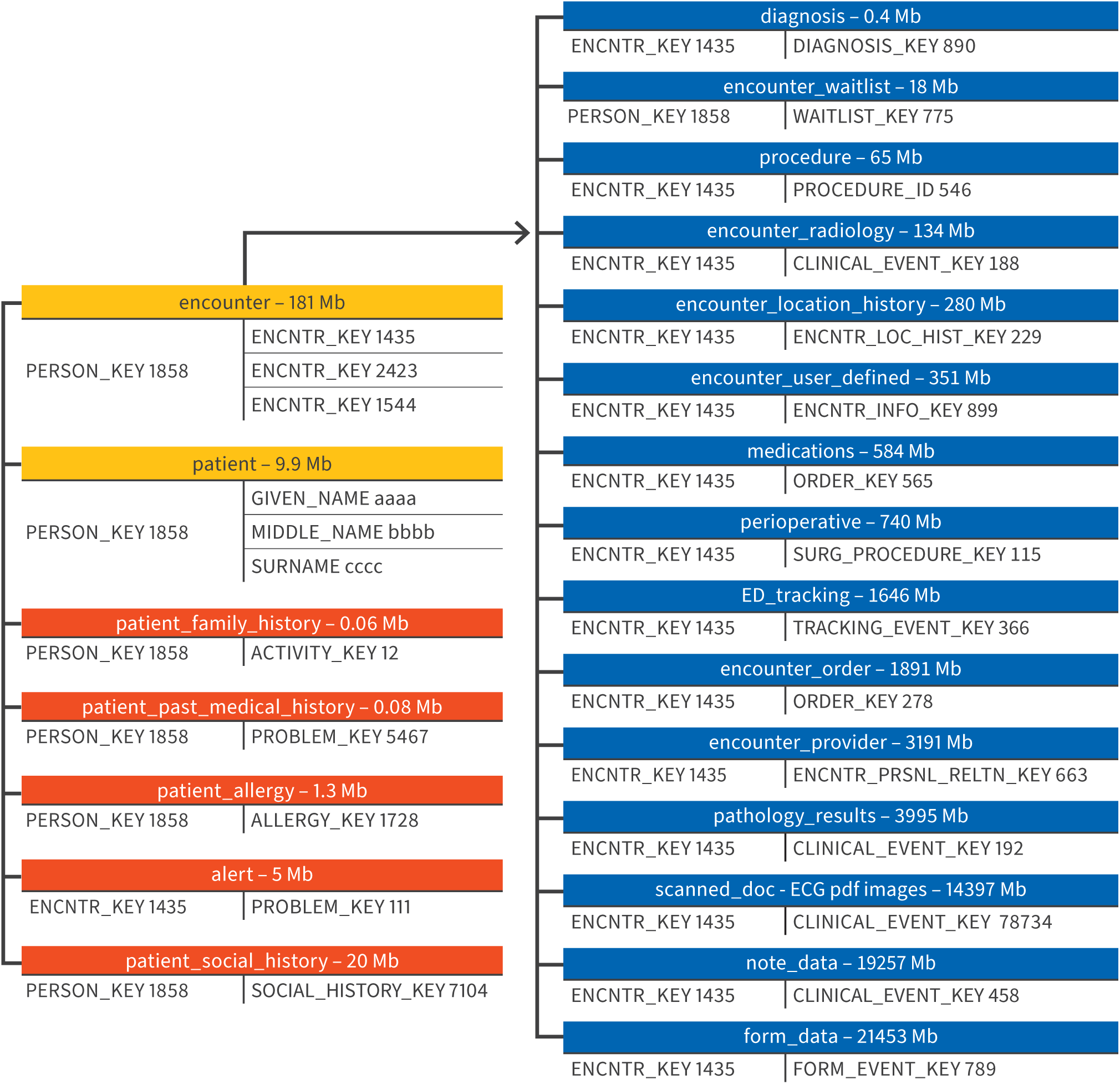
Visual representation of data tables extracted from Cerner Millenium eMR systems. Cerner eMR systems were extracted as data tables which are linked by encounter key. Data tables were linked by encounter key and include information about encounters, diagnoses (ICD10 and SNOMED-CT), pathology, forms (eg. ED triage assessments, medication forms, discharge letters), notes (eg. progress notes) and many more. Encounter level information were further linked to person level information by person key and included patient medical history and social history. The size of the raw file associated with each table is shown next to the table name. The data tables are listed in order of file size with the biggest files related to free-text information and scanned ECG images. The figure represents data extracted for a single 3 month time period (1/4/17-30/6/17).

### Outcome measures

eMR data extracts were processed and analysed using R (Version 4.0.0). Outcome measures included 1) examination of the relative contribution of individual inclusion criteria to the identification of eligible encounters, 2) associations between the use of diagnostic codes alone (ICD-10/SNOMED) *and* other inclusion criteria and 3) examination of consistency across LHDs and time. First, we developed a computational method to check the composition of inclusion criteria met by each encounter. Each encounter received a score between 0 and 7 indicating how many of the inclusion criteria were met. Any encounter with a score ≥1 was deemed eligible and included in the *index* cohort. Encounters that did not meet any of the inclusion criteria were assigned a value of ‘0’ and referred to as “*reference*” encounters (ie. they are not eligible encounters). Next, we examined the proportions of encounters identified using diagnostic codes alone *vs*. other inclusion criteria, and vice-versa and a correlation matrix was used to calculate the associations between each of the inclusion criteria. Finally, we examined consistency in the composition of inclusion criteria across time and local health districts.

## RESULTS

### Examination of the relative contribution of individual inclusion criteria to eligible encounters

The five-year extract (1/1/13-31/12/17) consisted of 802,742 eligible and 5,418,466 reference encounters. Out of the 802,742 eligible encounters, scanned ECG images (54.8% of encounters) and symptoms and possible ACS-related keywords in clinical documentation (41.4-64.0%) were used most often to identify presentations of possible ACS. Orders and investigations (27.3%) and procedures (1.4%), were less often present for identified presentations. Relevant ICD-10/SNOMED codes were present for 3.7% of identified encounters.

To further examine the composition of inclusion criteria in eligible encounters, UpSet plots [30] were used to represent the frequency of each inclusion criterion and the numbers of encounters that met each combination of inclusion criteria in 2017, the most recent data in our 5-year data extract. Figure 2 shows 185,414 eligible encounters in 2017 with similar findings as the total 5 year dataset. Individually, the presence of a scanned ECG image (72.0%), the presence of keywords captured in the presenting information in the ED triage form (60.0%) and the presence of keywords in the “Reason for visit” for the presentation (38.2%) identified the majority of eligible encounters. Orders and investigations (25.6%) and procedures (1.2%), were less often present for eligible encounters. Only 3.1% of encounters had the presence of a relevant diagnostic code (ICD-10 or SNOMED) for ACS. The presence of a cardiac monitoring form was not an informative criterion for identifying eligible encounters.

**Figure 2.**
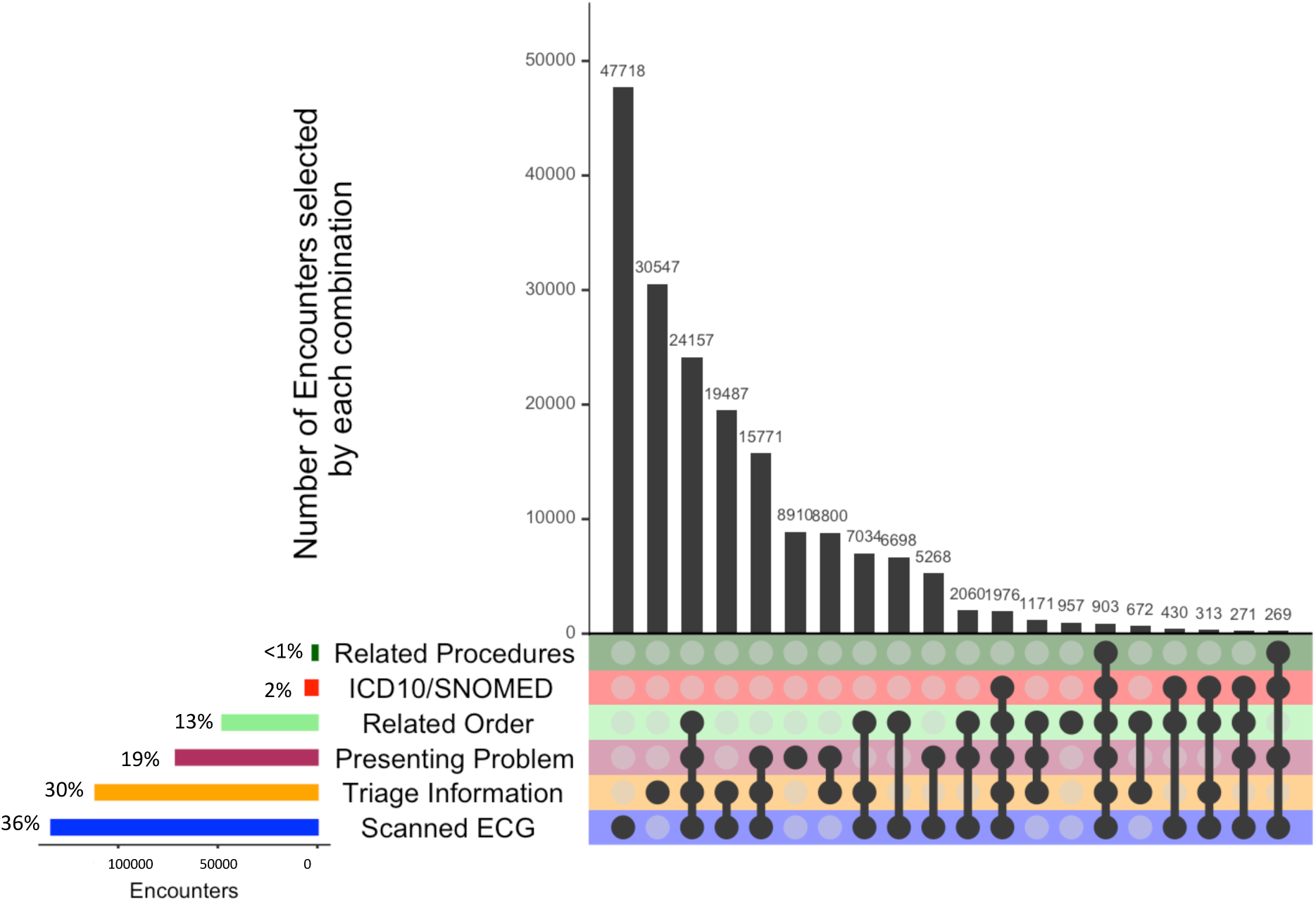
UpSet plot showing the number of eligible encounters meeting individual (bottom left hand side) and multiple inclusion criteria (right-hand side). This UpSet plot represents 317719 eligible encounters from Cerner information systems in two local health districts that met at least one of the study inclusion criteria in 2017. The bottom left-hand side represents the total number and percentage of encounters that met each inclusion criterion. Each inclusion criterion is represented as an independent group. The top right-hand side represents the number and percentage of encounters that met each combination of the inclusion criteria. Inclusion Criteria referred to: (1) *Presenting problem key match*: Keyword match in a free-text field for presenting information, (2) *Triage information key match*: The patient was assigned to a cardiac pathway mode of care or keyword match in the ED Triage descriptions, (3) *Related Order*: The existence of a cardiology-related order; (5) *Related Procedure*: the existence of cardiology-related procedure, (6) *ICD10/SNOMED:* The encounter had an SNOMED-CT or ICD10 code for Acute Myocardial Infarction (AMI) and (7) *Scanned ECG image:* The encounter had a scanned ECG imaget available. Inclusion Criterion (4), representing patients that had a cardiac monitoring form, was excluded as no encounters met this inclusion criterion.

We found 59 unique combinations of inclusion criteria met by encounters (Figure 2). The most frequent combinations were encounters that only had the presence of a scanned ECG image (26%; 47,514 of 185,414), encounters that only had a keyword match in triage information (18%; 33,865 of 185,414) and encounters that had the presence of an ECG, keyword match in triage information and a relevant order (12%; 22,160 of 185,414). Similar trends were seen in UpSet plots performed for the other years (2013-6; Supplementary Figure 1).

### Comparisons between diagnostic codes *and* other inclusion criteria

In the five-year extract, we examined the proportion of encounters that contained a relevant diagnostic code within each cohort of encounters met by each inclusion criterion. By design, diagnostic codes comprised a minor component of each individual inclusion criteria cohort (Figure 3). Note that 0.1% (858/802.742) of eligible encounters were identified using diagnostic codes alone. A correlation matrix examining associations between each of the inclusion criteria found a strong correlation between diagnostic code and relevant procedures (Pearson’s correlation coefficient= 0.9), with small to moderate associations with the other inclusion criteria (Supplementary Figure 2).

**Figure 3.**
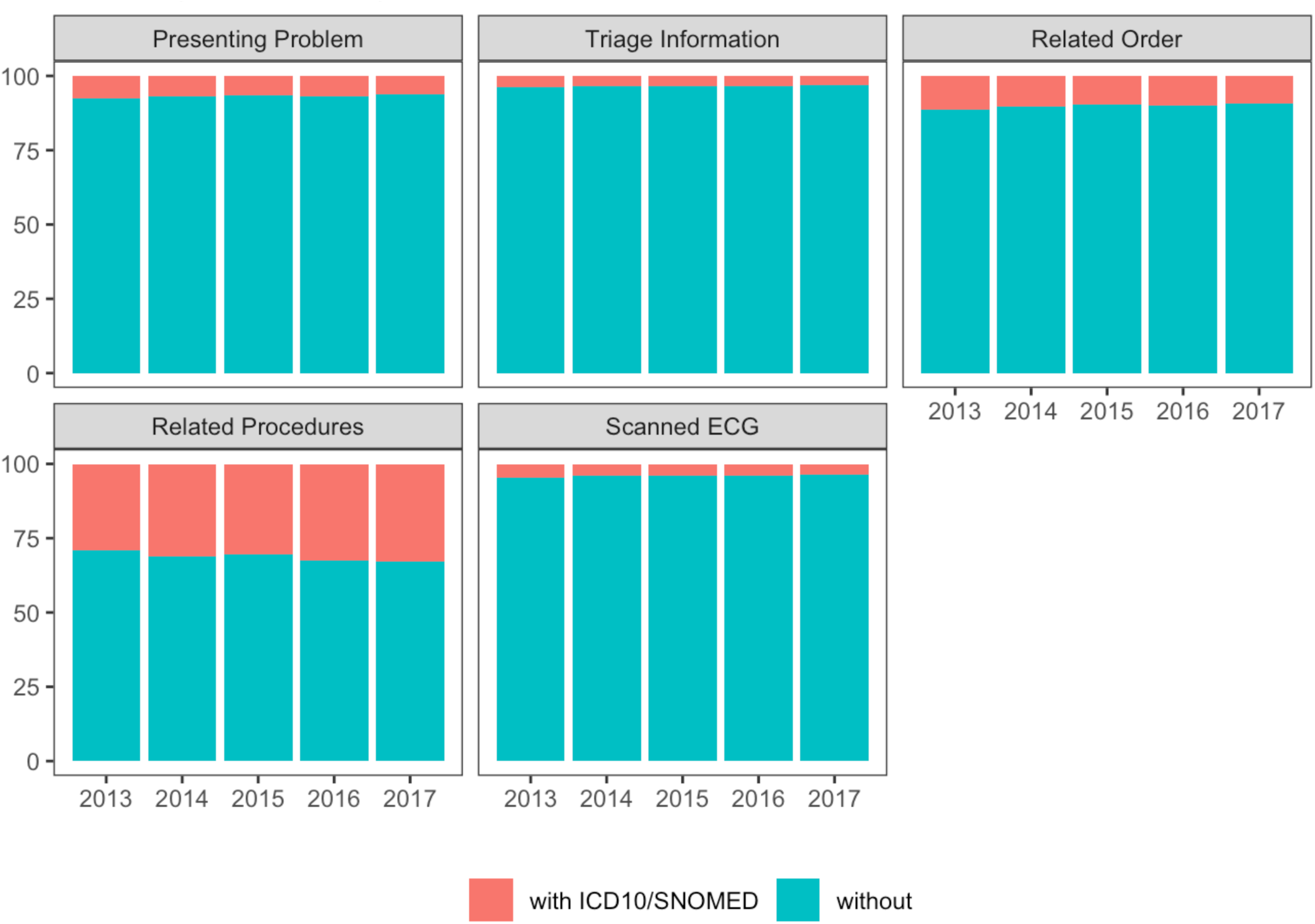
Percentage of eligible encounters with diagnostic codes (ICD10/SNOMED) for each inclusion criteria, by year.

### Consistency across local health districts and time

To examine consistency of the methodology, we examined the frequency of each inclusion criterion for each year in our 5 year extract (Table 2). Individually, the presence of a scanned ECG image (15-36%), the presence of keywords captured in the presenting information in the ED triage form (19-25%) and the presence of keywords in the “Reason for visit” for the presentation (30-40%) identified the majority of eligible encounters. Orders and investigations (13-17%) and procedures (<1%), were less often present for eligible encounters. 2-3% had the presence of a relevant diagnostic code (ICD-10 or SNOMED) for ACS. Similar trends were observed when the two LHDs were examined separately, with a diagnostic code being present in 2-3% of eligible encounters (Figure 4).

**Table 2.** Percentage of encounters that met each inclusion criteria in the 5 year cohort

| **Year** | **Presenting Problem,**<br>**n (%)** | **Triage Information,**<br>**n (%)** | **Related Order,**<br>**n (%)** | **Related Procedures,**<br>**n (%)** | **ICD10/**<br>**SNOMED,**<br>**n (%)** | **Scanned ECG**<br>**image,**<br>**n (%)** |
| --- | --- | --- | --- | --- | --- | --- |
| **2013**<br>**(n= 135511)** | 61529 (45.4) | 98045 (72.3) | 41629 (30.7) | 2296 (1.7) | 6240 (4.6) | 36902 (27.2) |
| **2014**<br>**(n= 143108)** | 65668 (45.8) | 99661 (69.6) | 42839 (29.9) | 2259 (1.6) | 5778 (4.0) | 46437 (32.5) |
| **2015**<br>**(n= 163008)** | 66428 (40.8) | 100048 (61.4) | 42734 (26.2) | 2099 (1.3) | 5613 (3.4) | 101363 (62.2) |
| **2016**<br>**(n= 175701)** | 68286 (38.9) | 105428 (60.0) | 45180 (25.7) | 2353 (1.3) | 5987 (3.4) | 121575 (69.2) |
| **2017**<br>**(n= 185414)** | 70878 (38.2) | 111275 (60.0) | 47566 (25.7) | 2243 (1.2) | 5768 (3.1) | 133429 (72.0) |

**Figure 4.**
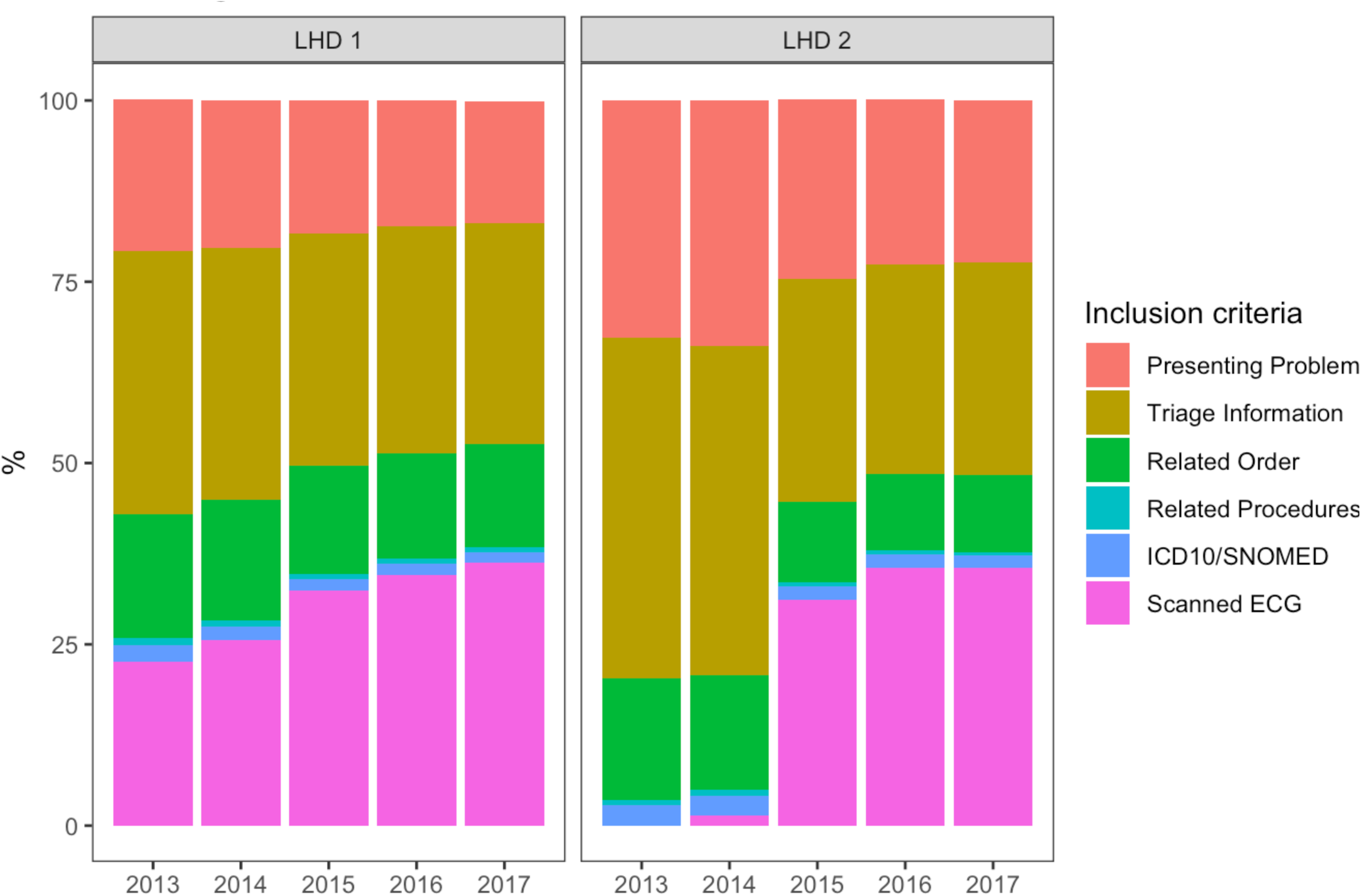
Percentage of total encounters in each local health district that met each inclusion criteria in a 5 year dataset. LHD 1 refers to Northern Sydney Local Health District and LHD 2 refers to Central Coast Local Health District.

## DISCUSSION

Our study demonstrates that informatics approaches combining structured eMR data, such as orders for pathology testing, investigations and diagnostic codes, with symptom and keyword text mining in narrative free-text during the data extraction process *within* eMR systems, can create high fidelity clinical-defined patient cohorts. This inclusive approach to eMR data extraction enables eMR-derived cohorts to be readily created and updated as new conditions/diseases emerge and clinical definitions are updated [31], as well as the extraction of clinically-relevant information enabling future validation studies of diagnostic and procedure codes, which are essential for secondary use of eMR data [9, 27]. This is particularly significant for diagnoses which have a diverse range of presenting problems (eg. ACS, mental illness, sepsis) [32].The use of diagnostic codes alone during this process would likely have led to relevant patients not being captured [33] as evidenced by the finding that <1% of eligible encounters contained an ACS-related diagnostic code [27].

Our work builds on earlier studies combining structured and unstructured free-text eMR data for identifying cohorts *after* data has been extracted from eMR systems, demonstrating that diagnostic codes alone are insufficient for disease case detection. Penz *et al* found that ICD-9 and current procedure terminology (CPT) codes identified less than 11% of the cases in a study of detecting adverse events related to central venous catheters, while natural language processing methods achieved a sensitivity of 0.72 and specificity of 0.80 [34]. Similar findings have been observed for detecting colorectal cancer cases [10]. Our study findings demonstrate that similar approaches for identifying cohorts are required *within* eMR systems themselves, which have also been demonstrated by large collaborative research networks (eg. PCORnet, eMERGE, ODHSI) [35].

The generalisable methodology described in this study is essential for curation of high-fidelity clinical data from eMR systems enabling continuous, routine monitoring and reporting on the quality of care and outcomes for an unselected cohort of patients across large health care treatment and referral networks or populations, as well as providing clinical decision-making guidance. Monitoring and reporting could be performed against agreed care standards and benchmarked outcomes for specific conditions, which would supplant the need for developing and maintaining labour intensive, condition-specific population-based observational cohort studies and clinical registries.

The overarching strength of the research is the liberal nature of the data extraction process enabling future validation work critical for secondary use of eMR data, flexibility for creating new eMR-derived cohorts as clinical definitions and guidelines get updated as well as being able to identify and extract information on patients based on presenting symptoms and investigations, rather than diagnostic code. To our knowledge, this is also the first time that clinically-relevant information for diagnosing ACS has been collated from eMR data extracts (Supplementary Table 1). Limitations of the research include that the study period was restricted to extracted data from one information system (Cerner Millenium), chosen as it was the main eMR system, from only two of the fifteen local health districts in New South Wales. Nevertheless the principles of this robust methodology can be applied to any eMR data extraction process and generalised to other diseases/conditions. Extending data extraction processes across health jurisdictions and for other conditions will enable further validation of the methodology.

## Conclusion

This paper demonstrates that clinically-defined eMR cohorts created using a broad strategy utilising structured and unstructured free-text, are likely to identify relevant cohorts of patients and enable critical validation work required for secondary use of eMR data.

## Data Availability

The data reported in this paper is patient data owned by the Health Organisation and is not publically available.

## Acknowledgments

This project was supported by the Sydney Informatics Hub, funded by the University of Sydney. We would also like to thank Ms Seven Guney and Mr Matthew Strasiotto for their assistance on this project and A/Prof Adam Dunn for critical feedback on the manuscript.

## Legends for Supplementary Figures

**Supplementary Figure 1.**
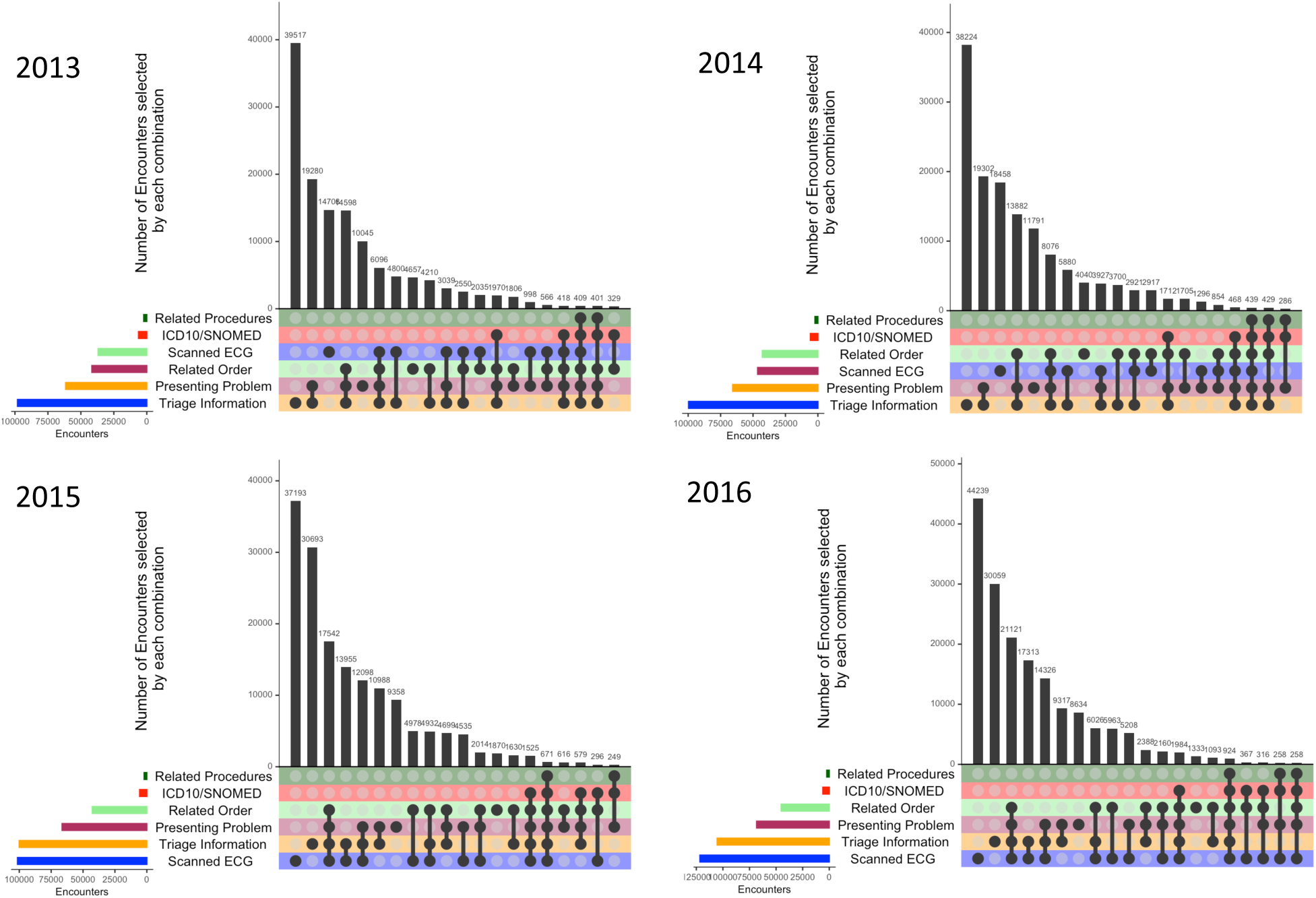
UpSet plot showing the number of eligible encounters meeting individual (bottom left hand side) and multiple inclusion criteria (right-hand side) in 2013-6. This UpSet plot represents eligible encounters from Cerner information systems in two local health districts that met at least one of the study inclusion criteria in 2013 (n=135,511), 2014 (n=143,108), 2015 (n=163,008) and 2016 (n=185,414). The bottom left-hand side represents the total number and percentage of encounters that met each inclusion criterion. Each inclusion criterion is represented as an independent group. The top right-hand side represents the number and percentage of encounters that met each combination of the inclusion criteria. Inclusion Criteria referred to: (1) *Presenting problem key match*: Keyword match in a free-text field for presenting information, (2) *Triage information key match*: The patient was assigned to a cardiac pathway mode of care or keyword match in the ED Triage descriptions, (3) *Related Order*: The existence of a cardiology-related order; (5) *Related Procedure*: the existence of cardiology-related procedure, (6) *ICD10/SNOMED:* The encounter had an SNOMED-CT or ICD10 code for Acute Myocardial Infarction (AMI) and (7) *Scanned ECG image:* The encounter had a scanned ECG report available. Inclusion Criterion (4), representing patients that had a cardiac monitoring form, was excluded as no encounters met this inclusion criterion.

**Supplementary Figure 2.**
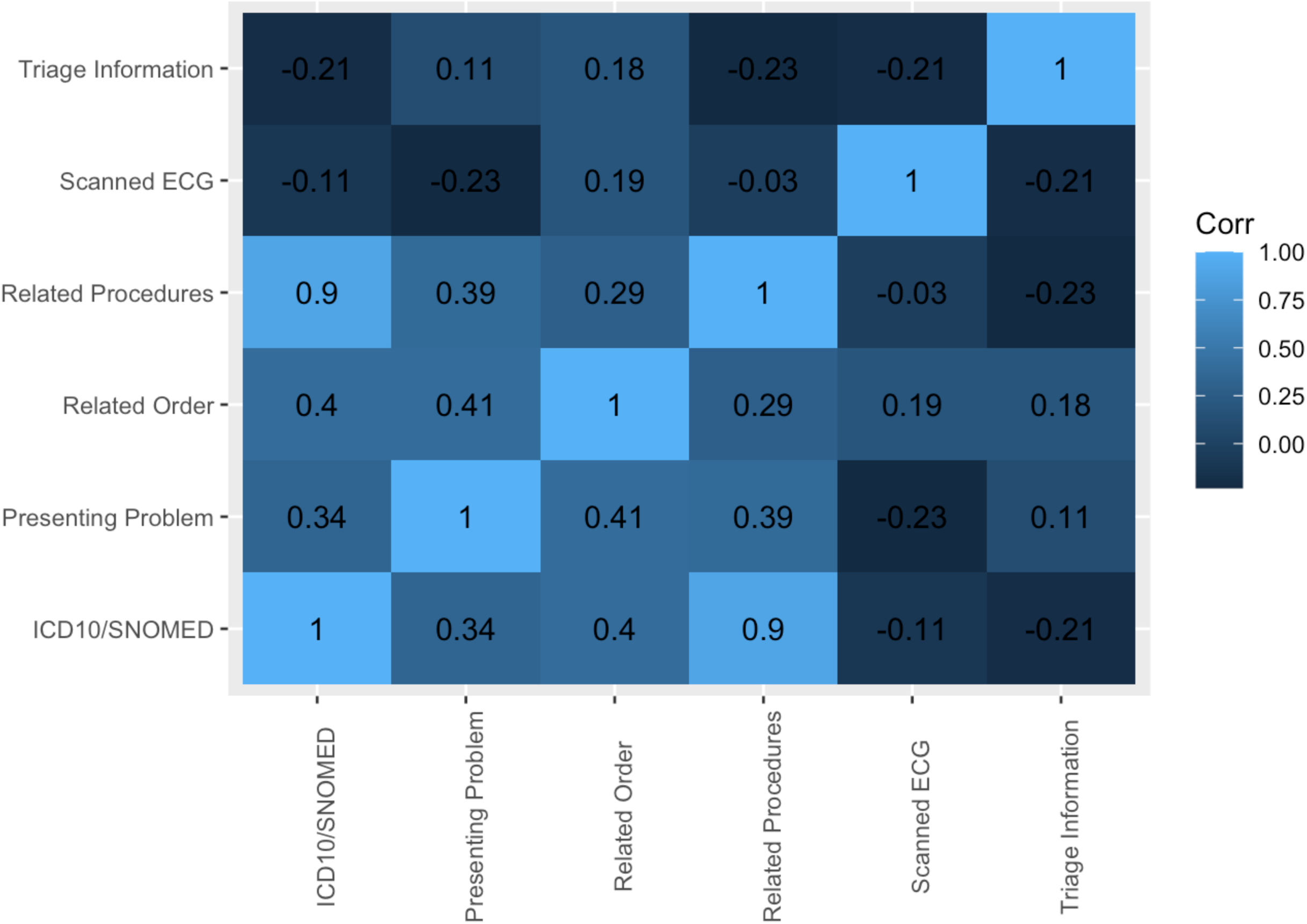
Correlation matrix examining associations between inclusion criteria

## Supplementary Information

### Supplementary Information Part 1

The following provides a list of search terms which forms part of inclusion criteria (1): “pain chest”| “pain,chest”| “pain-chest”| “chest pain”| “shortness of breath”| “sob”| “dizziness”| “vomiting”| “syncope”| “syncopal”| “presyncope”| “weakness”| “nausea”| “unwell” | “loc”| “cardiac arrest”| “nstemi”| “stemi”| “angio”| “angiogram”| “cor/angio”| “palpitation”| “salami”| “cabg”| “ami”| “coronary artery bypass graft”| “etami”| “heart attack”| “cath lab”| “cath”| “ohca”| “out of hospital arrest”| “out of hospital cardiac arrest”| “stent”| “fatigue”| “weakness”| “ventricular tachycardia”| “ventricular fibrillation”| “vt”| “vf”| “dyspnoea”| “chest tightness”

The following provides a list of search terms for inclusion criteria (2):

“ashen” | “chest pain”| “chest pains” | “chestpain”| “cp”| “chest tightness”| “clamminess”| “diaphoresis” | “diaphoretic”| “dizziness”| “dizzines” | “dizzy”| “dyspnoea”| “etami”| “fatigue”| “ingestion”| “indigestion”| “lightheadness”| “light headedness” | “lightheaded”| “light headed”| “light-headed”| “loc”| “loss of consciousness”| “nausea”| “pale”| “palpitations”| “palpatations”| “palpations”| “palpitation”| “palpitaion”| “palpatation”| “shortness of breath”| “short of breath”| “sob”| “stemi”| “nstemi”| “sweaty”| “sweats”| “syncopal”| “syncope”| “syncopy”| “syncople”| “vomiting”| “vomiting”| “vomitting”| “vomting”| “weakness”| “ohca”| “vt”| “ventricular tachycardia”| “vf”| “ventricular fibrillation”| “clammy”| “presyncope”| “presyncopal”| “chest heaviness”| “epigastric pain”| “arm heaviness”| “failed thrombolysis”| “thrombolysis”

### Supplementary Information Part 2

#### Approaches to assessing data quality

Data quality was assessed for completeness, accuracy and consistency [1].

##### Completeness

Completeness was assessed in several ways. Firstly, the percentage of missing values was ascertained for each data element. If data elements could be logically aggregated, then we examined the percentage of records with sufficient data to calculate an indicator or characteristic of interest (Eg. number of patients that had received a percutaneous coronary intervention). Ascertainment completeness, defined as the percentage of eligible cases present, was first compared by checking the face value of relevant characteristics with local clinicians. This was vital to develop confidence that all data elements for a required indicator had been identified and correctly extracted from the eMR. We also assessed the presence of duplicate records (Eg. diagnoses which were recorded twice), duplicate events (eg. a single procedure documented twice) and the absence of records (eg. All analytes which were measured in a single assay should have results recorded).

##### Accuracy

In the absence of a gold standard for assessing accuracy, or determining the true value of each record, we assessed validity, reliability, and as a theoretical step, the internal consistency of the encounters.

##### Validity (internal and external)

Internal validity was defined as the closeness of agreement between a data value and a plausible value. For pathology results, results were assessed against reference ranges. To verify time stamps, we checked that all time stamps fell within a patient’s admission and discharge time.

External validity was determined by checking distributions of variables and outliers, and comparing differences between our study, SPEED-EXTRACT and independent data sources published in similar clinical populations such as CONCORDANCE, a contemporary Australian observational registry that describes the management and outcomes in patients with ACS [2, 3] and another observational study of consecutive patients presenting with a ST-elevation myocardial infarction field triaged directly to the cardiac catheterisation laboratory within NSLHD [4]. Differences in results between SPEED-EXTRACT and published studies [2-4] were used to guide investigations as to whether data elements were missed during the data extraction process or characteristics were insufficiently captured. For example, smoking status can be collected as a structured field (eg. ex-, current- or non-smoker), however this was only filled in <1% of presentations, and tended to be collected in free-text clinical documentation.

##### Reliability

We examined relative uniformity in distributions of data elements across local health districts included in SPEED-EXTRACT (i.e., reproducibility) and temporal stability in the data extract across years at the same sites.

#### Operational framework

##### Data management and security

The SPEED-EXTRACT project brought together expertise across local health districts, university and government agencies (ie. NSW Ministry of Health, eHealth NSW). In doing so, it navigated existing governance processes and created new processes to enable this work to occur across traditional organisational and professional silos. Information from the eMR was extracted and de-identified on secure servers within the local health district. The de-identified data was then securely transferred to the University Research Data Store server for storage and analyses. The University Research Data Store and the High Performance Computing cluster are University facilities located in Tier 3 data centres in Sydney with an Information Security Management system that conforms to the requirements of ISO27001:2013 international standard [5] and all relevant NSW laws, regulations and statutory requirements. Discrepancies between identifiable eMR data extracted within the local health district and de-identified eMR data received by the University were checked by designated employee(s) of the local health district as an additional data quality measure to those described above. All code developed on the de-identified data for the project was stored on a version control system.

##### Project governance

The project was directed by an Executive Committee comprising leaders in cardiology and digital health, population health and data science experts and representatives from government agencies. The Committee provided advice on content and scientific issues such as development of data definitions for computable phenotypes, understanding of local clinical workflow practices and interpretation of ACS quality and safety indicators. A number of subgroups devolved from the Executive Committee including a Data and Analytics Team and a Publication Committee. The Data and Analytics Team comprised software engineers, data analysts, data scientists and analytics translators and used agile project methodology to address the clinical questions set by the Executive Committee.

**Supplementary Table 1.** Data availability in SPEED-EXTRACT compared to existing cardiovascular eMR studies.

| Data availability | SPEED-EXTRACT | CALIBER<br>[6] | CANHEART<br>[7] | CHERRY<br>[8] |
| --- | --- | --- | --- | --- |
| Presenting symptoms | ✓ | X | X | X |
| ECG | ✓ | X | X | X |
| Troponin | ✓ | X | X | X |
| Clinical documentation | ✓ | X | X | X |
| Current and new medications | ✓ | X | X | X |
| Procedures | Codes and details | Codes only | Codes only | Codes only |
| Diagnoses | Codes and clinician diagnoses | Codes only | Codes only | Codes only |
| Clinical outcomes (In-hospital mortality, intensive care admission, length of stay, readmission rates) | ✓ | ✓ | ✓ | ✓ |
| Linkage to existing administrative datasets | Future work, readily able<br>to be<br>performed | ✓ | ✓ | ✓ |

## References

1. Casey, J.A., et al., Using Electronic Health Records for Population Health Research: A Review of Methods and Applications. Annu Rev Public Health, 2016. 37: p. 61–81.

2. Haendel, M.A., C.G. Chute, and P.N. Robinson, Classification, Ontology, and Precision Medicine. N Engl J Med, 2018. 379(15): p. 1452–1462.

3. Devine, E.B., et al., Automating Electronic Clinical Data Capture for Quality Improvement and Research: The CERTAIN Validation Project of Real World Evidence. EGEMS (Wash DC), 2018. 6(1): p. 8.

4. De Coster, C., et al., Identifying priorities in methodological research using ICD-9-CM and ICD-10 administrative data: report from an international consortium. BMC Health Serv Res, 2006. 6: p. 77.

5. Johnson, E.K. and C.P. Nelson, Values and pitfalls of the use of administrative databases for outcomes assessment. J Urol, 2013. 190(1): p. 17–8.

6. Manuel, D.G., L.C. Rosella, and T.A. Stukel, Importance of accurately identifying disease in studies using electronic health records. BMJ, 2010. 341: p. c4226.

7. Shivade, C., et al., A review of approaches to identifying patient phenotype cohorts using electronic health records. J Am Med Inform Assoc, 2014. 21(2): p. 221–30.

8. Colborn, K.L., et al., Identification of urinary tract infections using electronic health record data. Am J Infect Control, 2019. 47(4): p. 371–375.

9. Botsis, T.H.G, Chen, F; Weng, C., Secondary use of EHR: Data quality issues and informatics opportunities. Summit on Translational Bioinformatics, 2010: p. 1–5.

10. Xu, H., et al., Extracting and integrating data from entire electronic health records for detecting colorectal cancer cases. AMIA Annu Symp Proc, 2011. 2011: p. 1564–72.

11. McDonald, H.I., et al., Methodological challenges when carrying out research on CKD and AKI using routine electronic health records. Kidney Int, 2016. 90(5): p. 943–949.

12. Al Sallakh, M.A., et al., Defining asthma and assessing asthma outcomes using electronic health record data: a systematic scoping review. Eur Respir J, 2017. 49(6).

13. Ingram, W.M., et al., Defining Major Depressive Disorder Cohorts Using the EHR: Multiple Phenotypes Based on ICD-9 Codes and Medication Orders. Neurol Psychiatry Brain Res, 2020. 36: p. 18–26.

14. Holve, E., C. Segal, and M. Hamilton Lopez, Opportunities and challenges for comparative effectiveness research (CER) with Electronic Clinical Data: a perspective from the EDM forum. Med Care, 2012. 50 Suppl: p. S11–8.

15. Sun, W., et al., Data Processing and Text Mining Technologies on Electronic Medical Records: A Review. J Healthc Eng, 2018. 2018: p. 4302425.

16. Abhyankar, S., et al., Combining structured and unstructured data to identify a cohort of ICU patients who received dialysis. J Am Med Inform Assoc, 2014. 21(5): p. 801–7.

17. Carroll, R.J., et al., Portability of an algorithm to identify rheumatoid arthritis in electronic health records. J Am Med Inform Assoc, 2012. 19(e1): p. e162–9.

18. Kreuzthaler, M., S. Schulz, and A. Berghold, Secondary use of electronic health records for building cohort studies through top-down information extraction. J Biomed Inform, 2015. 53: p. 188–95.

19. Fernandez-Breis, J.T., et al., Leveraging electronic healthcare record standards and semantic web technologies for the identification of patient cohorts. J Am Med Inform Assoc, 2013. 20(e2): p. e288–96.

20. Virani, S.S., et al., The use of structured data elements to identify ASCVD patients with statin-associated side effects: Insights from the Department of Veterans Affairs. J Clin Lipidol, 2019. 13(5): p. 797–803 e1.

21. Ford, E., et al., Extracting information from the text of electronic medical records to improve case detection: a systematic review. J Am Med Inform Assoc, 2016. 23(5): p. 1007–15.

22. Burns, E.M., et al., Systematic review of discharge coding accuracy. J Public Health (Oxf), 2012. 34(1): p. 138–48.

23. Horsky, J., E.A. Drucker, and H.Z. Ramelson, Accuracy and Completeness of Clinical Coding Using ICD-10 for Ambulatory Visits. AMIA Annu Symp Proc, 2017. 2017: p. 912–920.

24. Healthstats, NSW. http://www.healthstats.nsw.gov.au/Indicator/dem_pop_age/dem_pop_lhn_snap 2020 x1/2/20].

25. Benchimol, E.I., et al., The REporting of studies Conducted using Observational Routinely-collected health Data (RECORD) statement. PLoS Med, 2015. 12(10): p. e1001885.

26. Aschard, H., et al., Evidence for large-scale gene-by-smoking interaction effects on pulmonary function. Int J Epidemiol, 2017. 46(3): p. 894–904.

27. van Walraven, C., C. Bennett, and A.J. Forster, Administrative database research infrequently used validated diagnostic or procedural codes. J Clin Epidemiol, 2011. 64(10): p. 1054–9.

28. Hemingway, H., et al., Big data from electronic health records for early and late translational cardiovascular research: challenges and potential. Eur Heart J, 2018. 39(16): p. 1481–1495.

29. Rubbo, B., et al., Use of electronic health records to ascertain, validate and phenotype acute myocardial infarction: A systematic review and recommendations. Int J Cardiol, 2015. 187: p. 705–11.

30. Conway, J.R., A. Lex, and N. Gehlenborg, UpSetR: an R package for the visualization of intersecting sets and their properties. Bioinformatics, 2017. 33(18): p. 2938–2940.

31. Pendergrass, S.A. and D.C. Crawford, Using Electronic Health Records To Generate Phenotypes For Research. Curr Protoc Hum Genet, 2019. 100(1): p. e80.

32. Jackson, R.G., et al., Natural language processing to extract symptoms of severe mental illness from clinical text: the Clinical Record Interactive Search Comprehensive Data Extraction (CRIS-CODE) project. BMJ Open, 2017. 7(1): p. e012012.

33. van Walraven, C. and P. Austin, Administrative database research has unique characteristics that can risk biased results. J Clin Epidemiol, 2012. 65(2): p. 126–31.

34. Penz, J.F., A.B. Wilcox, and J.F. Hurdle, Automated identification of adverse events related to central venous catheters. J Biomed Inform, 2007. 40(2): p. 174–82.

35. Rasmussen, L.V., et al., Considerations for Improving the Portability of Electronic Health Record-Based Phenotype Algorithms. AMIA Annu Symp Proc, 2019. 2019: p. 755–764.

## References

1. https://rethinkingclinicaltrials.org/. NIH Collaboratory Living Textbook of Pragmatic Living Trials 2020 1/2/20].

2. Aliprandi-Costa, B., et al., The design and rationale of the Australian Cooperative National Registry of Acute Coronary care, Guideline Adherence and Clinical Events (CONCORDANCE). Heart Lung Circ, 2013. 22(7): p. 533–41.

3. Khan, E., et al., Differences in management and outcomes for men and women with ST-elevation myocardial infarction. Med J Aust, 2018. 209(3): p. 118–123.

4. Vernon, S.T., et al., Increasing proportion of ST elevation myocardial infarction patients with coronary atherosclerosis poorly explained by standard modifiable risk factors. Eur J Prev Cardiol, 2017. 24(17): p. 1824–1830.

5. https://www.iso.org/standard/54534.html. ISO/IEC 27001:2013. 2019.

6. Denaxas, S.C., et al., Data resource profile: cardiovascular disease research using linked bespoke studies and electronic health records (CALIBER). Int J Epidemiol, 2012. 41(6): p. 1625–38.

7. Tu, J.V., et al., The Cardiovascular Health in Ambulatory Care Research Team (CANHEART): using big data to measure and improve cardiovascular health and healthcare services. Circ Cardiovasc Qual Outcomes, 2015. 8(2): p. 204–12.

8. Lin, H., et al., Using big data to improve cardiovascular care and outcomes in China: a protocol for the CHinese Electronic health Records Research in Yinzhou (CHERRY) Study. BMJ Open, 2018. 8(2): p. e019698.

